# Female Saudi nursing students’ perspectives on pursuing a career in home healthcare: A qualitative study

**DOI:** 10.1101/2023.01.10.23284389

**Authors:** Wafa Hamad Almegewly, Savvato Karavasileiadou, Taghreed Samel Alotaibi

## Abstract

**Background:** While the number of patients requiring home healthcare in Saudi Arabia is increasing, there is insufficient data on what attracts nurses to work in this field.

**Objective:** This qualitative study investigates how nursing students practicing in home healthcare perceive it as a future career.

**Methods:** A purposive sample was used to recruit fourth-year nursing students enrolled in an older adult nursing care course and trained in home healthcare settings. Data were gathered using five focus groups of five students each (a total of 25 students) and analyzed using thematic analysis.

**Results:** It was found that the majority of students did not perceive home healthcare as a viable career option compared to working in a hospital. They vacillated due to the nature of the work, safety concerns, working demands, invariability of health cases, and lack of opportunities for professional development. Nevertheless, some nursing students were amenable to pursuing a career in home healthcare due to the less working hours, sense of autonomy, and the ability to provide holistic care and educate patients and their families.

**Conclusion:** Population awareness programs are needed to overcome cultural barriers, increase student motivation and ultimately bolster the number of certified nurses working in home healthcare.

## 1 Background

Home healthcare refers to the process of caring for patients suffering from acute, terminal, and chronic health conditions at the patient’s home or designated care facilities in the community (1). Home healthcare also provides various community services, including preventive, nursing, and psychological care and support (6). This practice is unlike hospital-based services with advanced facilities that provide specific care for acute and chronic conditions; home healthcare primarily involves families, caregivers, and patients (2). In Saudi Arabia, home healthcare programs were started at the Green Crescent Hospital in 1980, followed by King Faisal Specialist Hospital and Research Centre in 1991, and King Fahad National Guard Hospital (KFNGH) in 1995 (3). Since then, many governmental and private healthcare organizations have implemented similar programs and offered similar services across the country (5).

Home healthcare nursing has seen enhancements and growth as a result of demographic changes such as patient age, chronic diseases, and healthcare access. For example, in the US, 70.5% of home healthcare users are older adults with multiple chronic health conditions who need continuous monitoring (4), similar to the situation in Saudi Arabia (5,6). In addition, the rapid adoption of new technologies, such as telemedicine (7) and telephonic nursing follow-ups (8), has enhanced access to home healthcare, minimized hospital stay length, and improved quality of life (9).

Despite global developments in home healthcare, this specialized field still faces major nursing shortages (7). One of the considerable challenges in Saudi Arabia is a shortage of nurses in this field due to poor management, lack of suitable infrastructure, cultural barriers, low salary, inter-professional coordination and cooperation, and job dissatisfaction (10,11). Other contributing factors include a lack of home healthcare equipment, insufficient security measures, and quality indicators (12). These issues contribute to nurses’ stress and burnout (10).

Several studies have focused on the challenges nurses face in home healthcare, while others have addressed factors that make the practice desirable. A sense of autonomy contributes to work satisfaction and nurses’ desire to pursue home healthcare as a career (13). The sense of autonomy refers to individual freedom and the ability to make independent choices (14,15). Other factors include favourable employment conditions, having the space and time to offer quality care, work flexibility, and support from supervisors (16,17)

Although the literature has provided valuable information on the challenges and benefits of home care nursing work, it has only focused on the working nurse’s perspective. No research has been conducted on prospective nursing students in the home healthcare profession. There is a need for qualitative studies which provide an in-depth analysis of all these factors to encourage the recruitment and retention of homecare nurses.

Saudi nursing students especially lack home healthcare skills and knowledge. More detrimental is that this specialization is not often incorporated into the nursing curriculum of most nursing colleges (18). Most nursing students’ clinical training generally focuses on hospital-based practice for all nursing competencies. Both governmental and private nursing schools fail to include home healthcare nursing in their list of mandatory clinical practices (19). Consequently, nursing students have limited knowledge of home healthcare and nursing duties compared to the key focus on the hospital-based learning experience. Moreover, the Saudi healthcare system is highly reliant on nurses (8). In addition, home healthcare is primarily run by nurses with fewer direct interventions from physicians. Further exacerbating these challenges are the nursing shortage, a lack of nursing student interest in community work, and a growing number of older adults needing home healthcare. The present study addresses some of these challenges by describing how nursing students in home healthcare perceive the specialization as a potential future career.

Research questions:

1) What are nursing students’ perceptions toward working in home healthcare?

2) What factors motivate nursing students to work in home-based healthcare settings?

## 2 Methods

### 2.1 Research design

A qualitative descriptive phenomenological study approach (20) was used to describe nursing students’ different experiences and perceptions toward home healthcare as a future career. An in-depth investigation was also conducted into the factors that motivate nursing students to work in home-based healthcare settings. The Consolidated Criteria for Reporting Qualitative Research (COREQ) (21) was used to ensure the study was satisfactorily reported.

### 2.2 Research setting and sample

The study was implemented at a Saudi government university for females in the College of Nursing, Riyadh, Saudi Arabia. A purposive sample of undergraduate female nursing students was selected according to the following inclusion criteria. They were (a) female nursing students enrolled in a baccalaureate nursing program. The nursing students must be (b) in their fourth year of study, enrolled in a course on nursing care for older adults, and trained in older adult home healthcare as part of the course curriculum. The selected college was incorporating home healthcare training into its curriculum for the first time. The total number of eligible participants was N = 144; the collected sample size was n = 25. This was based on the “code saturation” as no more new frequent codes were noticed. Also, “information power” was used for better understanding of the discussed issues which focused on the rich information retrieved from the participants regardless their small number.

### 2.3 Data collection process

All students who met the inclusion criteria were invited to participate in a focus group interview by the academic student affairs unit. The research team developed a topic guide based on existing literature to investigate the nursing students’ overall experience in nursing home healthcare. This guide focused on students’ feelings and opinions of home healthcare nursing and the benefits, challenges, and receptiveness to working in the specialization. Props were used to encourage student participation and provide clearer explanations and examples. Experts on the home healthcare topic guide were consulted to ensure the validity of the questions.

Ultimately, 25 students participated in five focus groups of five students each. The focus groups emphasized interaction and discussion among the participants and described the different views in a safe, respectful environment. The groups were facilitated by the first author, conducted in Arabic, and lasted for approximately one hour. As the first author was a faculty member, many precautions were taken to minimize coercion problems. Firstly, the selected participants were not taught by the first author. Secondly, participants were informed that participation was voluntary and that their grades would not be affected. All focus groups were conducted in one of the College of Nursing meeting rooms.

Audio of all five focus groups was recorded and password protected to safeguard participant confidentiality. Only research investigators had access to this information. The interviews were transcribed verbatim into English, verified using the audio recordings, and reviewed for accuracy after the first and third authors assessed them. Since, the first and third authors were bilingual, they checked the accuracy of the verbatim translation to ensure the credibility and validity of the results after translation.

### 2.4 Ethical consideration

Ethical approval for the study was obtained from the selected university (ref no:19-0199). Informed consent was obtained from all participants by the research team, who explained that they could withdraw from the study at any time. All study-related information was explained to the participants by the research team, who had no direct relationship with them to avoid any incidence of coercion. Students were also reassured that their marks would not be affected. Selected identification letters and numbers were assigned to each student to promote the confidentiality and anonymity of the participants.

### 2.5 Data analysis

Inductive thematic analysis was used to analyse the data (22). First, the researchers read the transcripts several times to familiarize themselves with the data and ensure its accuracy. Second, initial notes and codes were generated. Third, patterns and similarities across the data were identified, and themes were developed. Fourth, the main themes and subthemes were formulated. Finally, themes and supporting data from transcript extracts or quotes were outlined.

## 3 Results

Three central themes emerged. These themes focused on the benefits of working in home healthcare nursing, the challenges of working in home healthcare nursing, and working in home healthcare vs. hospital. Respondents also noted that the nature of home healthcare nursing work differs from work in hospital settings. Furthermore, students discussed their opinions on working as nurses in home healthcare and whether they would choose home healthcare as a future career path.

### 3.1 Benefits of working in home healthcare nursing

The students mentioned several benefits related to working in home healthcare nursing during the focus groups, including independence, and closeness and trust.

#### 3.1.1 Independence at work

Independence at work identified as main them emerge from students focus group interviews. As from frequent students’ answers home healthcare nurses are independent in their decision-making, with a high feeling of autonomy and self-efficacy in delivering complete care to older adults in their homes, which includes evaluation, intervention, and education. Several responses from the student to describe dependency at work the following student describe sense of autonomy by decision making:

> *“The good side of home healthcare nursing is that a nurse is able to make decisions and focus on one patient*.*”* (C2)

The other students achieve independence by dealing with the patient and their family one-to-one:

> *“I only have a patient and caregivers, which increases my independence. I give medicine and act on the treatment plan for the patient*.*”* (A4)

> *“I am alone with the patient, not with too much advanced medical equipment, and that makes me feel more independent*.*”* (C1)

Also, some students commented in their abilities to do an environmental assessment along with educating the patients and his or her household members:

> *“For the first time, I was able to do an environmental assessment*.*”* (E3)

> *“I got the chance to educate the patient as well as the caregivers*.*”* (A4)

#### 3.1.2 Closeness and trust

The students understood the importance of the homecare nurses’ ability to establish trust with their patients and families. Students discussed the great esteem in which patients and their families hold home healthcare nurses. Furthermore, being an Arabic-proficient Saudi student also helped foster trust and strengthened the relationships with patients and their families:

> *“The trust between the patient and the nurse in home healthcare nursing is higher than in the hospital*.*”* (B3)

Further students describe presence of an Arabic-proficient Saudi student on fostering trust and strengthening relationships:

> *“Some patients were very glad that I’m Saudi and I took care of them. Their words were encouraging [*…*]. I mean yes, they (patients) need Saudi nurses because we can understand them*.*”* (D4)

> *“I remember when I first entered the patient’s house, they [patient’s family] thought that I’m a foreigner, but when I spoke in Arabic, they were shocked that I’m Saudi*.*”* (C4)

The nurses also bonded with their patients because of frequent and long-term visits. One of the nurses said that “*if the patient’s case resulted in deterioration or death, [she] would be very sad as the patient was viewed as part of [her] family*.” (A2)

Many students felt confident performing basic nursing skills and assessments, as the environment was conducive to learning. This was often contrary to their experience in the hospitals, where some patients underestimated the knowledge and skills of the nursing students:

> *“In home healthcare, I feel that I’m closer to my patients, I can do patients’ assessment from different angles like social, psychological, environmental, and physical*.*”* (B4)

### 3.2 Challenges working in home healthcare nursing

The students indicated several difficulties with training in home healthcare. Most importantly was a lack of preparation, a difficult and demanding setting, and an uncomfortable atmosphere.

#### 3.2.1 Unanticipated situations

Students frequently expressed concerns about facing unanticipated issues in home healthcare and lack of training in how to deal with aggressive and violent patients and family caregiver neglection while the incident occurs in their homes:

> *“We were supposed to have an introductory course on home care nursing and about the student’s challenges*.*”* (A3)

Some students have questioned the skills they learned during the course and felt that they were not sufficiently prepared to deal with older adults and violent cases:

> *“One day the patient was yelling at me and screaming. I got scared. The college didn’t prepare us how to deal with such cases like violence or harassment or how to comfort patient*.*”* (E4)

#### 3.2.2 Complex and demanding setting

Students reported that working in home healthcare requires a particular personality type and an affinity for managing and communicating with older adult patients.

> *“Home healthcare nurses mostly deal with elderly patients and honestly, I don’t know really how to deal with them*.*”* (C3)

> *“I was excited to teach the patients, but at the same time, I was anxious because of responsibility on my shoulders as I’m the only Saudi female who’s giving them the information*.*”* (B4)

> *“The nurse acting as a social worker, psychologist. She was doing multiple roles at once”* (E5).

In addition, some students felt that while the sense of autonomy in home healthcare is high, this puts more pressure on them to do all the care work independently, with minimal help from others.

> *“The nurse there […] is overwhelmed with the working load because she was taking over all the health healthcare team roles like being the doctor, educator, phlebotomist, and the physiotherapist*.*”* (C2)

> *“Tele-nurse should be implemented at the home healthcare so the patients can ask or receive any information. In terms of the nursing training, I would recommend less paperwork and more hands-on*.*”* (C4)

### 3.3 Home healthcare vs. hospital

The primary student perception was that they would prefer to work in a hospital rather than a home healthcare setting. Furthermore, they indicated how different the nursing jobs performed in the two environments were.

#### 3.3.1 Working nature

Students highlighted the lower workload in home healthcare, which consists primarily of basic tasks, when compared to hospital tasks. One commentator, for example, said the following to describe nursing work as routine work and not requiring specialized care:

> *“Most of the cases were stable and didn’t require a complex care, […] it was a routine work*.*”* (E2)

Furthermore, some students felt that the workload in home healthcare is low compared to hospitals, as there are fewer complications during a home healthcare visit and no need for further care interventions over and above what is expected:

> *“I might consider working in home healthcare in the future because the workload is light and most of the cases are stable*.*”* (C3)

> *“At home healthcare, the working load is 8 hours compared to 12 hours at the hospital*.*”* (E1)

Students also felt that the home healthcare nursing tasks were too routine and limited to patient vital sign assessments, which are not as beneficial as hospital training.

> *“The home healthcare will not add that much to my professional development. There I’ll spend the whole day checking the vital signs, and this is very routine*.*”* (D3)

Safety concerns were a key reason for some students opting for a career in hospitals. They felt that there are safety measures in place at hospitals, while home healthcare teams are put at risk when they travel to unsafe locations to visit their patients:

> *“The hospital is more secure and safer than home healthcare. Here you are responsible for your own safety*.*”* (A1)

> *“I went to remote areas. The houses were in poor conditions and located in unsafe neighbourhoods*.*”* (A1)

#### 3.3.2 Professional growth

Most interviewed students disregarded home healthcare nursing as a career option and preferred hospitals for various reasons. First, the students focused on a stimulating specialization where they could develop their clinical skills and have more opportunities for professional advancements, such as in the emergency department or intensive care unit.

Some students stated hospital nurses are skilful and describe home care nurses because as limited to communication skills with older people:

> *“I think hospital nurses are skilful as they practice on a daily basis. But in-home nursing, nurses’ skills are limited as they can only follow one patient’s condition*.*”* (B2)

Students perceived hospital nurses’ work descriptions as more structured than those of home healthcare nurses, where patient-cantered care is the primary focus:

> *“At the hospital, everyone knows his/her responsibilities*.*”* (B4)

## 4 Discussion

This qualitative study provided valuable insights into the perceived benefits and challenges of home healthcare from the nursing students’ perspective. Some students found home healthcare appealing because of patients’ hospitality and gratitude, as a sense of belonging is important for nurses’ work motivation (23).

Furthermore, many participants reported positive experiences as they learned to overcome their fears, be more independent, and become more patient-cantered. This personal development is facilitated by the nature of home healthcare, which has a one-to-one nurse-patient ratio. These circumstances nurture closer practitioner-patient relationships and enable strong interpersonal bonds to form. A study in Canada showed that nursing students gained valuable insights into their patients but also developed confidence in their abilities and took more initiative with their self-directed learning. Consequently, they experienced greater autonomy and independence over time (24). Calma et al. (2019) demonstrated the importance of characteristics particular to home healthcare nurses, such as personality, capability, self-confidence, sociability, communication skills, and autonomy (25). Some of these factors were common to the present study, especially autonomy.

Participants indicated feelings of independence in home healthcare. They could communicate with patients in a common language, which helped them understand their patient’s condition and provide better feedback and education. Anecdotal evidence shows that 70% of nurses in the Saudi healthcare system do not speak Arabic or come from different cultures (9,10). This may limit their ability to communicate efficiently with patients. Therefore, policymakers, healthcare organizations, educators, and healthcare professionals must adopt practices that help attract and retain Saudi nurses working in home healthcare (26).

Some students have experienced difficulties handling the holistic approach required in home healthcare nursing. According to Nabolsi et al. (2012), nurses’ responsibilities and roles, such as records maintenance and patient treatment, can vary substantially (27). Some discussions with the participants in the present study regarding their workload indicated that home nursing was a highly demanding job compared to other professions (28).

The current study demonstrated that some students felt uncomfortable or unsafe in the home nursing environment, negatively affecting their perspective on the specialization as a potential career. Research also shows that home healthcare clinics are considered an unsafe and sometimes violent work environment (29).

Accessibility of the patient homes also appears to be a challenge globally (30). The present study found that some students could not travel to a dwelling for work or were uncomfortable with traveling long distances. While the participants were familiar with the hospital location and routes, home healthcare settings are often not easily accessible or safe to approach. Thus, it is essential to ensure the accessibility and safety of home healthcare nursing for students and healthcare professionals.

Many participants were uncertain about choosing home healthcare as a career because it was perceived as a complex and demanding setting. Most nursing students prefer to work in hospitals due to better opportunities to develop professionally and apply critical thinking skills. This finding is in line with existing research (27,31,32). In the Netherlands, Van Iersel et al. (2018) found that only 5.4% of first-year nursing students would consider working in home healthcare compared to 72% who would rather work in a general hospital (33).

Research also finds that nursing students perceived home healthcare as physically demanding, unappealing, unchallenging (34), undistinguished, and insufficiently equipped to handle the needs of the patients (35). However, there were nursing students who felt fulfilled by the experience and expressed a desire to pursue a career in home healthcare. Other students were uncertain of what career specialization to pursue. This can be attributed to a lack of experience or knowledge of the potential nursing career paths (27). The nursing curriculum and clinical practices design substantially impact students’ preferences and career choices (36). Similarly, students’ career preferences are influenced by the programs and modules they undertake at the university level (37). Universities and educators should consider this when designing university curricula to support nursing students in career planning.

The students sampled in the present study wanted a better-structured training program. Some were shocked by the lack of preparation or introductory lectures. Some key areas for improvement in nursing education include high-quality clinical placements for exposure and skill practice. In addition, targeted educational interventions should be included in the curriculum (38). A well-designed curriculum that includes a preparatory introductory course can help ensure professional competence and quality of home healthcare. It is important to note that clinical practice can shape student perceptions (39). Clinical placements in community care should offer students the opportunity for inter-professional collaboration, a chance to deal with the complexities of home-based patients, and engagement in an advanced care role (28). Moreover, students should be supported by specialized staff who can work across universities and placement settings (40).

A well-designed preparatory course must be included in the curriculum. Continuous medical education, including academic activities to increase home healthcare knowledge, is paramount. These student development aspects can be complemented by participation in conferences and research. Furthermore, media, workshops, seminars, and symposia can promote home healthcare knowledge and awareness. The need for extracurricular activities is apparent in a study from the Eastern Province of Saudi Arabia, which found that hospital-based experience was the primary source of information for health professionals (11).

Many studies have highlighted mentors’ contributions to healthcare education (37). Bjork et al. (2014) highlighted that the preceptor and clinical instructor play crucial roles in assessing training progress (41). Similarly, Bisholt et al. (2014) revealed that home healthcare behaviors are substantially influenced by clinical instructor’s attitudes, indicating that her presence plays a role in building student’s confidence (36). Research has found that the pedagogical atmosphere fostered by the relationship with supervisors was a factor in shaping first-year nursing students’ perceptions of older adult care (39).

### 4.1 Strengths and limitations

This study makes a valuable contribution to the existing literature as no qualitative studies have been conducted on home healthcare nursing in the Middle East and Saudi Arabia. This study provides a novel perspective on how student nurses perceive careers in home nursing. Furthermore, this study outlines the benefits and challenges of working in homecare nursing. A crucial aspect of home healthcare was identified in the interviews: the patient experience. When a nurse from the same background, speaking the same language, was treating them, patients and their families felt more comfortable—patient comfort is vital to patient care.

This study has some limitations. First, only a small sample from a single government institution was used, which may not be representative of all nursing students in Saudi Arabia. Consequently, the results may have limited generalizability. Second, only interviews were used for data collection. As such, mixed methods studies may be beneficial in future research. Future studies should engage in multi-methods research. Broader focus elements should be included, such as the experience of nurses of both genders, all grades, and from variable institutions (public and governmental). Further insights might be gained by comparing the findings of these studies with similar healthcare studies from other regions and settings.

### 4.2 Implications

Implementing population awareness programs on the role of home healthcare is needed. This will help set realistic expectations and overcome cultural barriers and prejudices in patients and their families. Better-equipped home healthcare, clear practitioner responsibilities, and continuous onsite personnel and educational support can improve nursing students’ career satisfaction and nurture a sense of professional development. Furthermore, early progressive involvement of nursing students in home healthcare during their academic curriculum—starting from the first year—will familiarize them with the benefits of working in home healthcare. Clinical placement environments should be educationally enriched with equipment and support from multidisciplinary teams and practitioners. This will provide more opportunities for professional development. Lastly, educators should provide individualized student mentorship via interviews or group meetings. This can aid in understanding their motivation, educational needs, and other concerns. Educators must realize their role in influencing student career choices.

## 5 Conclusion

This study provides valuable insights into Saudi nursing students’ current attitudes and perceptions toward home healthcare as a career option. Notably, the majority of students did not perceive home healthcare as a viable career option compared to working in hospitals. Their hesitation was largely due to the nature of the work, safety issues, lack of experiential diversity, and lack of opportunities for professional development. Regardless, some nursing students expressed an interest in home healthcare due to variable factors. These included limited working hours, a sense of autonomy, less demanding care, and the ability to educate patients and their families.

## Data Availability

Data cannot be shared publicly because of confidentiality of students

## 6 Acknowledgements

The authors express their gratitude to Princess Nourah bint Abdulrahman University Researchers Supporting Project number (PNURSP2023R312), Princess Nourah bint Abdulrahman University, Riyadh, Saudi Arabia.

## 7 Author contributions

Conceptualization, W.A and T.A; methodology, W.A, T.A and S.K; formal analysis, W.A AND T.A; review and editing, W.A, T.A and S.K.

